# Beneficial immune-modulatory effects of the N-163 strain of Aureobasidium pullulans-produced 1,3-1,6 Beta glucans in young boys with Duchenne muscular dystrophy: Results of an open-label, prospective, exploratory clinical study

**DOI:** 10.1101/2021.12.13.21267706

**Authors:** Kadalraja Raghavan, Vidyasagar Devaprasad Dedeepiya, Subramaniam Srinivasan, Subramanian Pushkala, Sudhakar Subramanian, Nobunao Ikewaki, Masaru Iwasaki, Rajappa Senthilkumar, Senthilkumar Preethy, Samuel JK Abraham

## Abstract

**Background:** This trial is to evaluate the effects of supplementation of Aureobasidium pullulans-N-163 strain produced 1,3-1,-6 beta glucan in young patients with Duchenne muscular dystrophy (DMD).

**Methods:** Twenty-seven male subjects aged 5-19 years with DMD were included, nine in the control arm and 18 in the treatment arm to receive N-163 beta glucan along with conventional therapies for 45 days. While performing the analysis, patients were stratified into: those not administered steroids (Steroid -ve) (Control, n=5; treatment, n=9), those administered steroids (Steroid +ve) (Control, n=4; treatment, n=9), which was not pre-specified.

**Results:** IL-6 showed a significant decrease in the N-163 Steroid -ve group, from 7.2 ± 1.2 pg/ml to ± 0.03 pg/ml. IL-13 decreased in both treatment groups—from 157.76 ± 148.68 pg/ml to 114.08 ± 81.5 pg/ml (N-163 Steroid -ve) and from 289.56 ± 232.88 pg/ml to 255.56 ± 214.13 pg/ml (N-163 Steroid +ve). TGF-β levels showed a significant decrease in the N-163 Steroid –ve group. Dystrophin levels increased by up to 32% in both treatment groups. Medical research council (MRC) grading showed slight improvement in muscle strength improvement in 12 out of 18 patients (67%) in the treatment group and four out of nine (44%) subjects in the control group.

**Conclusion:** Supplementation with the N-163 beta glucan food supplement produced beneficial effects: a significant decrease in inflammation and fibrosis markers, increase in serum dystrophin and slight improvement in muscle strength in DMD subjects over 45 days, thus making this a potential adjunct treatment for DMD after validation.

**Trial registration:** The study was registered in Clinical trials registry of India, CTRI/2021/05/033346. Registered on 5th May, 2021.

## Introduction

Duchenne muscular dystrophy (DMD) is a devastating X-linked neuromuscular disorder causing severe and progressive weakness of skeletal muscles, leading to loss of ambulation along with concomitant impairment of cardiac and respiratory muscles and early mortality. Mutations in the dystrophin gene, which cause total loss of the dystrophin protein [2], remain the major underlying mechanism. Loss of dystrophin leads to damage of the myofibres’ plasma membranes and distorts the structural stability of the plasma, leading to weakness in the myofibres. The weakened myofibres cannot withstand the contraction and relaxation cycles occurring during muscle function. The damage to the membrane releases the cytoplasmic contents, triggering the immune system and causing further muscle fibre damage, weakness and ultimately death [3]. A chronic proinflammatory state ensues, with neutrophil infiltration and macrophages’ phagocytosis of the degenerated tissue [3], preventing repair of the muscle damage, which otherwise occurs in a highly orchestrated manner for faster repair in other physiological conditions. The muscle is relatively immunologically privileged, with a low capacity to generate localized immune responses and thus having low rates of abscess and granuloma formation [3]. Therefore, it becomes essential to modulate the inflammation and immunity to resolve the chronic inflammatory state in therapeutic approaches to DMD. Steroid therapy is the most commonly employed immuno-modulatory treatment approach. However, side effects include weight gain, weak bones, high blood pressure and behaviour changes in addition to muscle weakness and atrophy in the long term [4,5]. Thus, there arises the need to develop strategies that will assist in immunomodulation with lesser side effects. Nutritional supplements are a potential option. Beside beta glucans yielding locomotor improvement in zebrafish models of DMD [6], a 1-3,1-6 beta glucan from the N-163 strain of the black yeast Aureobasidium pullulans has been reported to mitigate inflammation, evident by decreases in anti-inflammatory markers such as CD11b, serum ferritin, galectin-3 and fibrinogen. It also produces beneficial immuno-modulation via a decrease in the neutrophil-to-lymphocyte ratio (NLR) and an increase in the lymphocyte-to-CRP ratio (LCR) and leukocyte-to-CRP ratio (LeCR) in human healthy volunteers [7]. Mitigation of lipotoxicity-associated inflammatory cascades in a mouse study has also been reported [8]. Another study done in an animal model of non-alcoholic steatohepatitis (NASH) showed a decrease in liver inflammation and accumulation of F4/80+ cells (macrophages associated with inflammation) [9] in the liver. The present exploratory study is to evaluate the immunomodulatory efficacy of the N-163 strain of A. pullulans-produced beta 1-3,1-,6 glucan in comparison with a conventional therapeutic regimen in patients with DMD.

## Methods

This trial was an investigator-initiated, single-centre, randomized, open-label, prospective, comparative clinical study of patients with DMD. The study was conducted over 45 days. The two treatment arms included were:

***Treatment arm I***, control arm: Conventional treatment regimen comprising standard routine physiotherapy for joint mobility along with medications, viz., T. calcium and vit. D 1000 with or without deflazacort (steroid) 6mg to 24 mg.

***Treatment arm II***, intervention: One sachet of N-163 beta glucan (15g gel) once daily along with conventional treatment.

The stratification based on steroid was performed only during analysis of the data and was not pre-specified.

### Inclusion criteria

Male subjects with molecular diagnosis of DMD aged 6-18 years who were willing to participate in the study with written informed consent.

### Exclusion criteria

Patients with a previous (within the past 1 month) or concomitant participation in any other therapeutic trial; a known or suspected malignancy; any other chronic disease or clinically relevant limitation of renal, liver or heart function according to the discretion of the investigator.

### Investigations

The following tests were carried out after written consent was obtained from the study subjects.

### At baseline and at the end of the study (after 45 days)

- Background survey: gender, date of birth, age, habits, current medical history, medication, treatment, allergies (to drugs and food), regular use of food for specified health uses, functional foods, health foods, intake of foods rich in β-glucan foods containing beta-glucan and intake of immunity-boosting foods
- Medical history and physical measurements: height, weight, BMI, temperature
- Physiological examination: systolic blood pressure, diastolic blood pressure, pulse rate
- ECG
- Muscle strength test using MRC grading [10]
- Six-minute walk test (6MWT) [11]
- North Star Ambulatory Assessment (NSAA) [12]
- Blood sampling and investigations for the levels of IL-6, IL-13, TGF-β, creatinine kinase (CK), titin, haptoglobin, TNF-α, dystrophin, cystatin in the blood and myoglobin in the urine
- Subjects were contacted every week for drug compliance and recording of adverse effects, if any

### Study subjects = 28

The study was designed as an exploratory study, so there were two intervention conditions: one control and one test group. As the minimum number of participants required for statistical comparisons within and between intervention conditions is four per intervention condition, a total of 28 target study participants (10 in treatment arm I [control] group and 18 in treatment arm II [N-163]) were used. One patient was disqualified due to misrepresentation of diagnosis. The CONSORT flow diagram of the trial is shown in Figure 1.

**Figure 1:**
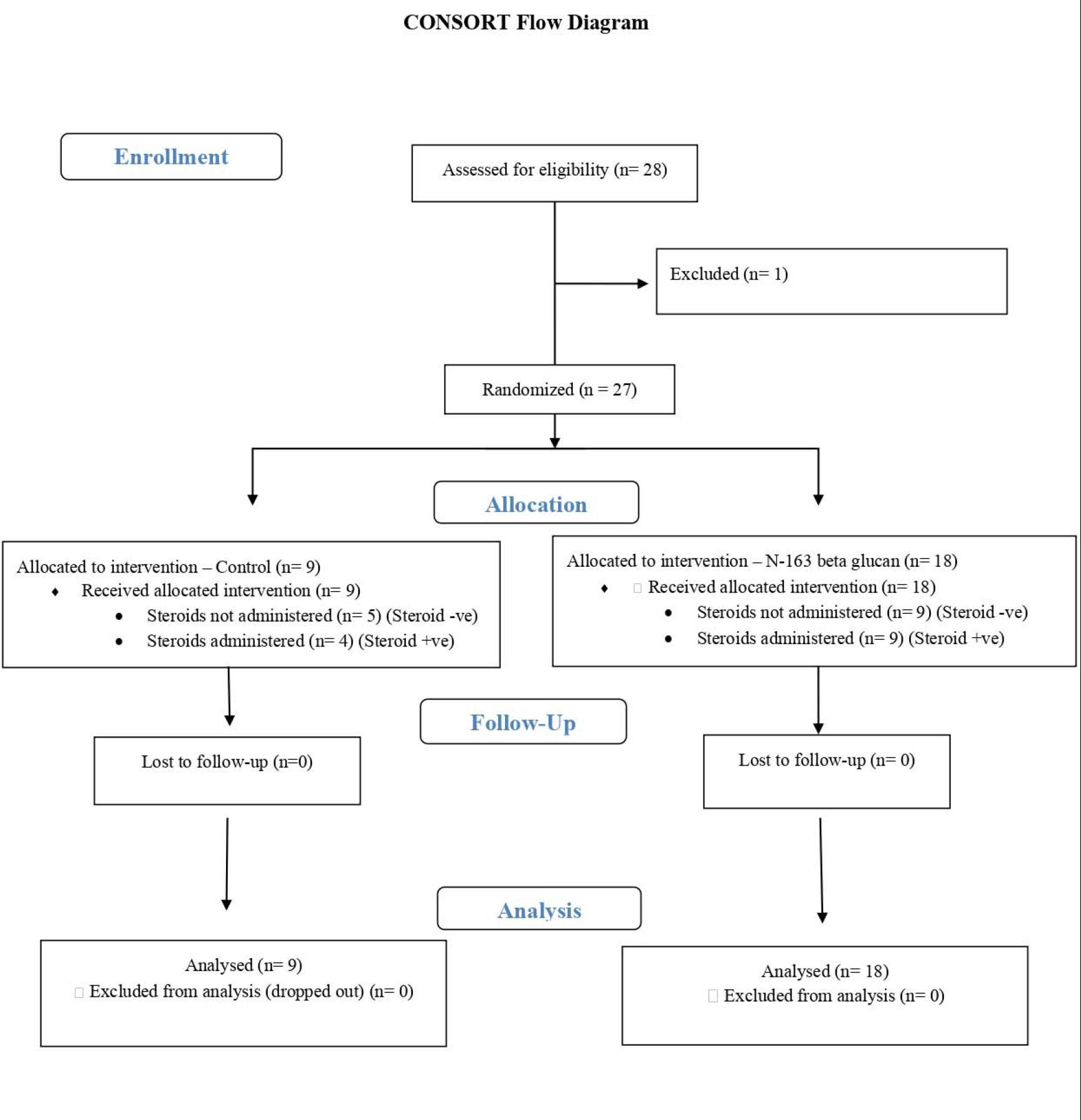
CONSORT flow diagram of the trial

### Selection of study subjects

Study investigators and other investigators included study subjects who had consented to participate in the study, met the selection criteria and not the exclusion criteria, and who were judged to have no problem participating in the study.

### Allocation of study subjects

The person in charge of the allocation, as specified in the study protocol, allocated the study subjects to the two groups by simple randomization (1:2).

### Primary outcome

Observation of changes in the levels of IL-6 in serum and myoglobin levels in urine from the baseline measured by ELISA.

### Secondary outcome

- Observation of changes in the levels of IL-13, TGF-β, CK, titin, dystrophin, haptoglobin and cystatin C in the serum measured by ELISA.
- Observation of changes in the muscle function tests
- Monitoring for adverse effects

### ELISA

The biomarkers were validated using sandwich enzyme immunoassay. ELISA kits were purchased from different manufacturers and the assay details along with sensitivity are listed in Table 2. ELISAs were performed according to manufacturer’s instructions and the serum was diluted to fall within the linear range of each respective assay.

**Table 1:**
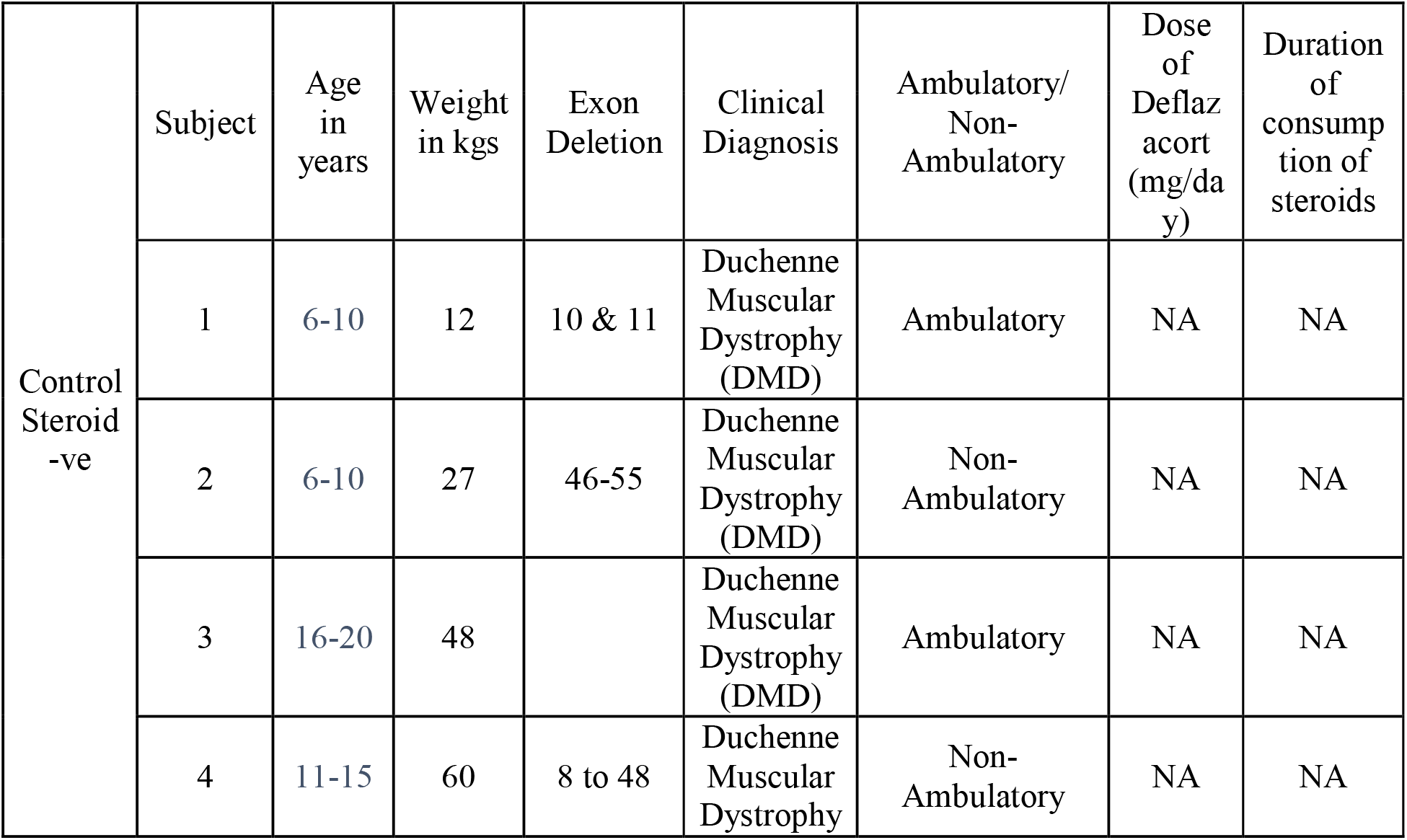
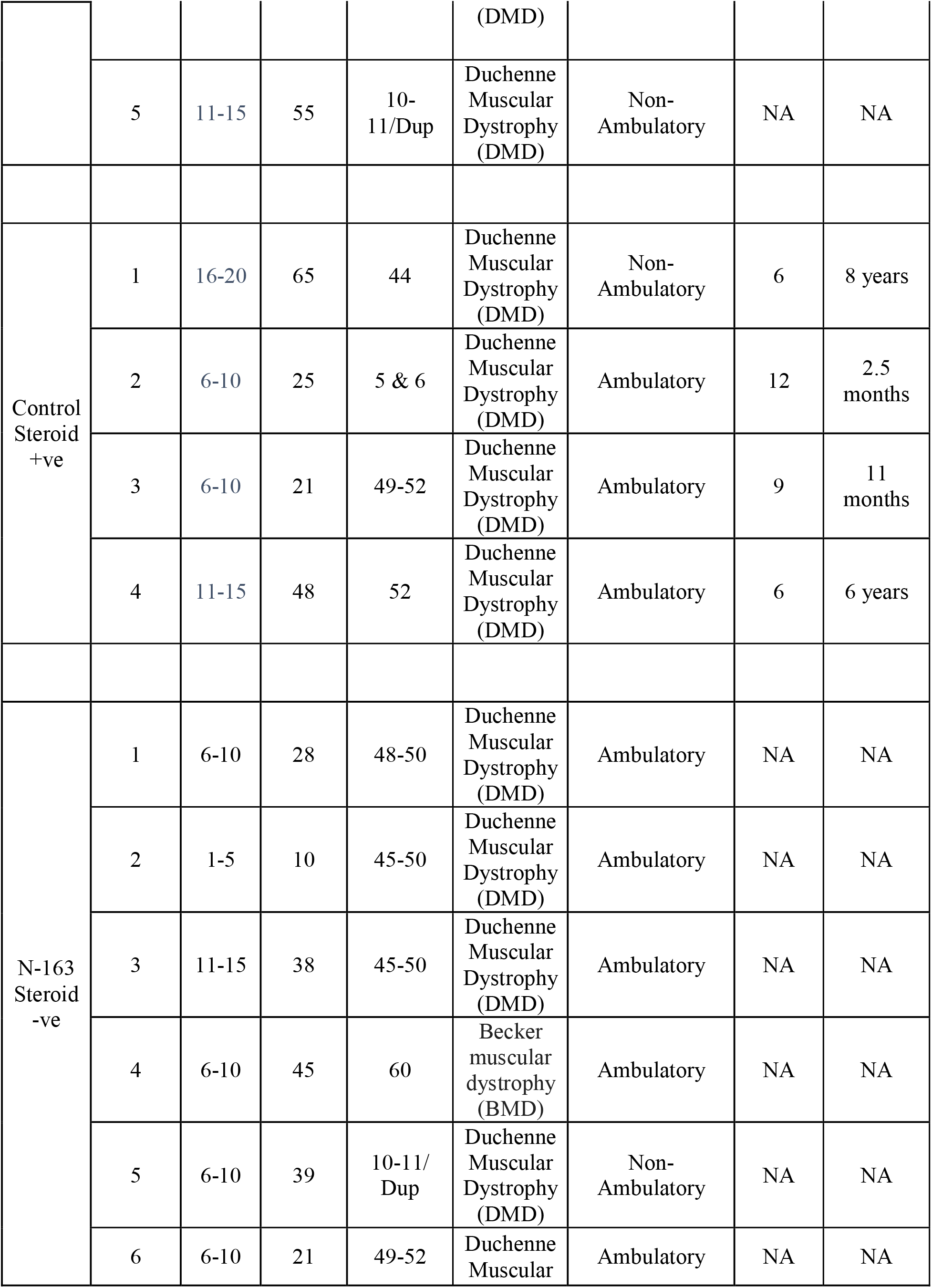
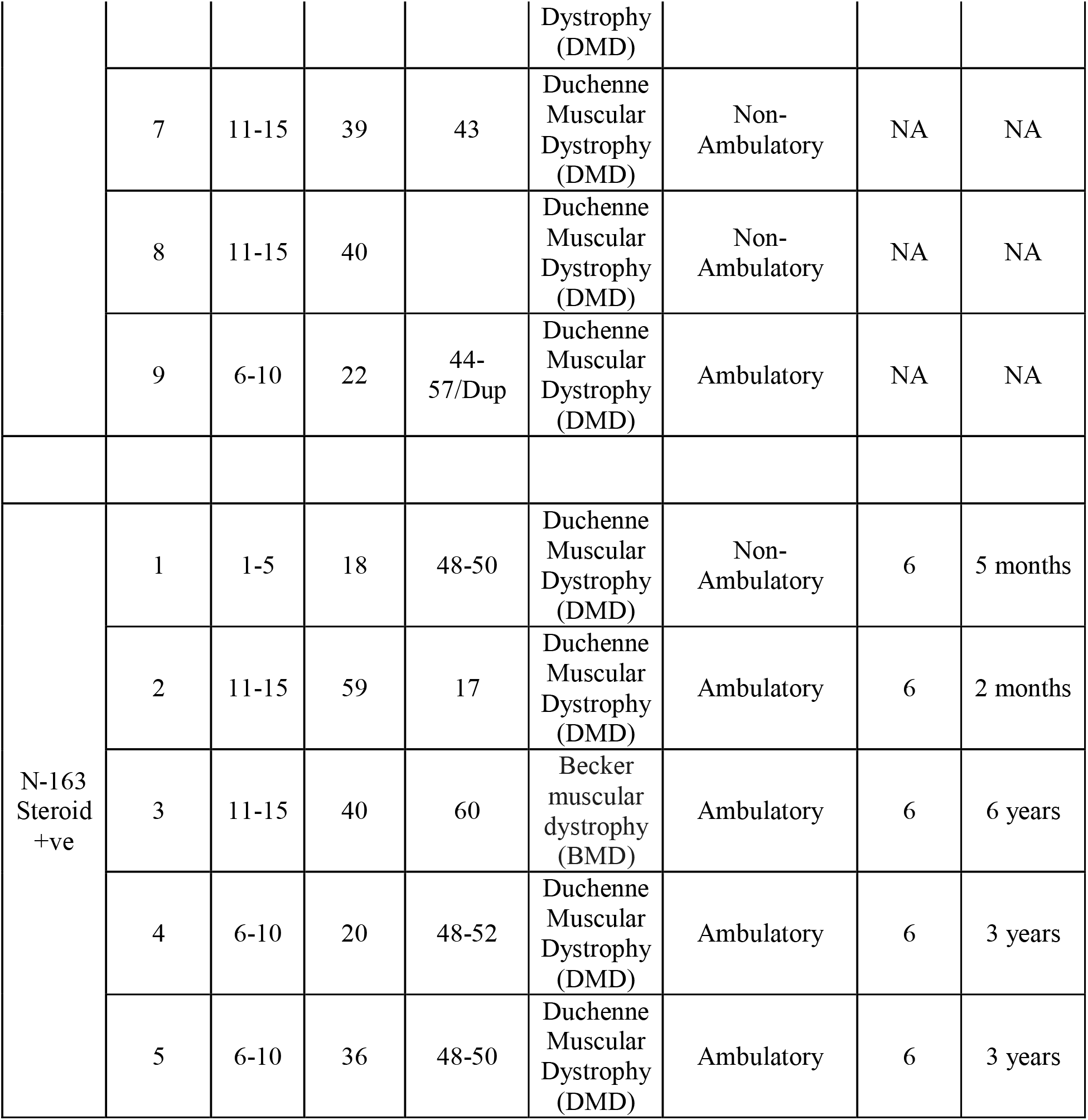
Demographics and baseline characteristics

**Table 2:** Details of the ELISA kits along with their sensitivity

| S.No | Human Antibody | Purchased from | Catalogue No. | Limit of detection/Sensitivity |
| --- | --- | --- | --- | --- |
| 1 | Interleukin-6 (IL6) | Seimens | 10995080 | 3.0 pg/mL |
| 2 | Myoglobin | Diagnostic Automation Inc | 1667-18 | 270 pg/mL |
| 3 | Dystrophin (DMD) | KINESISDx | K12-1446 | 0.042 ng/mL |
| 4 | Titin (TTN) | Mybiosource | MBS762344 | 0.094 ng/mL |
| 5 | Creatinine Kinase (CK) | Agappe | 51404001 | 2.0 U/L |
| 6 | IL-13 | Diaclone | 850080048 | 1.5 pg/mL |
| 7 | TGF-β | Diaclone | 650010096 | 8.6 pg/mL |
| 8 | Cystatin C | Siemens | E0001V | 0.06 μg/L |
| 9 | TNF-a | Siemens | LKNF1 | 6.23 pg/mL |
| 10 | Haptoglobin | Seimens | OSAV09 | 0.26 ng/mL |

### Statistical analysis

All data were analysed using Excel statistics package analysis (Microsoft Office Excel) and Origin2021b software. All 27 randomized participants were included in safety and efficacy analysis sets. All statistical tests were performed at a significance level of 0.05 without correction for multiple comparisons or multiple outcomes. Non-parametric tests such as Mann-Whitney U test and Kruskal-Wallis tests were used.

## Results

Twenty-eight patients were screened and 27 were randomized to control (n = 9) and treatment (n = 18). One patient was disqualified due to misrepresentation of diagnosis. The CONSORT flow diagram of the trial is shown in Figure 1.

Demographics are shown in Table 1.

The stratification of patients was as follows based on steroid administration during analysis:

Group I: Control group (n = 9);

A. Steroids not administered (n = 5) (Steroid -ve)
B. Steroids administered (n = 4) (Steroid +ve)

Group II: Treatment (N-163) group (n = 18);

A. Steroids not administered (n = 9) (Steroid -ve)
C. Steroids administered (n = 9) (Steroid +ve)

The mean ± SD age for the total study population was 11.18 ± 3.86 years (range 5-19 years). The mean age across the groups were Control (Steroid-ve) −14.25 years; Control (Steroid +ve) – 13.6 years; N-163 (Steroid -ve) – 9.6 years; N-163 (Steroid +ve) – 11.87 years. However the difference was not statistically significant (P= 0.316). The mean ± SD body weight was 35.59 ± 15.5 kgs (range = 10 to 65 kgs).

No adverse events were reported. No clinically significant changes from baseline data were observed on physical examination or in vital signs—temperature, blood pressure, oxygen saturation, pulse rate or ECG (data not shown).

### Biomarker levels

Levels are expressed as mean ± SD. In regard to the primary outcome, IL-6 showed decrease in both the control and the treatment groups. However, when the analysis was stratified for steroid administration, IL-6 showed the highest decrease in the N-163 Steroid -ve group, from a baseline value of 7.2 ± 1.2 pg/ml to 2.7 ± 0.03 pg/ ml post intervention and the difference was significant (p-value = 0.01) (Figure 2, 6).

**Figure 2:**
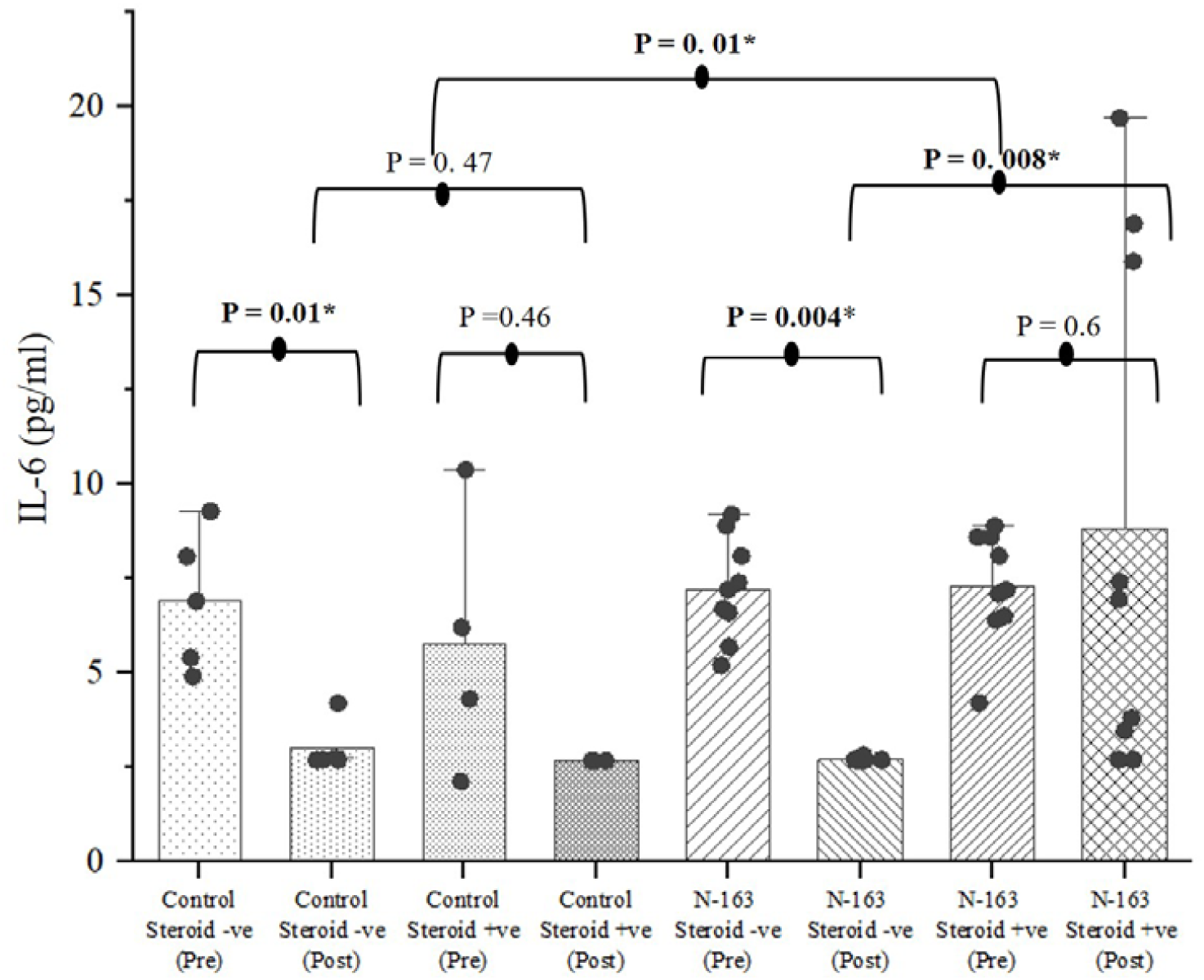
IL-6 showed the most significant decrease in the N-163 Steroid -ve group compared to other groups. (*p-value significance < 0.05)

IL-13 increased in both control groups—from 300.4 ± 114.5 pg/ml at baseline to 550.732 ± 107.95 pg/ml post-intervention in the Steroid -ve group and from 142 ±112.82 pg/ml at baseline to 263.5 ± 99.38 pg/ml post-intervention in the Steroid +ve group. It decreased in both the treatment groups—from 157.76 ± 148.68 pg/ml at baseline to 114.08 ± 81.5 pg/ml post-intervention and from 289.56 ± 232.88 pg/ml at baseline to 255.56 ± 214.13 pg/ml post-intervention. The difference was statistically significant (p-value = 0.006) (Figure 3, 6).

**Figure 3:**
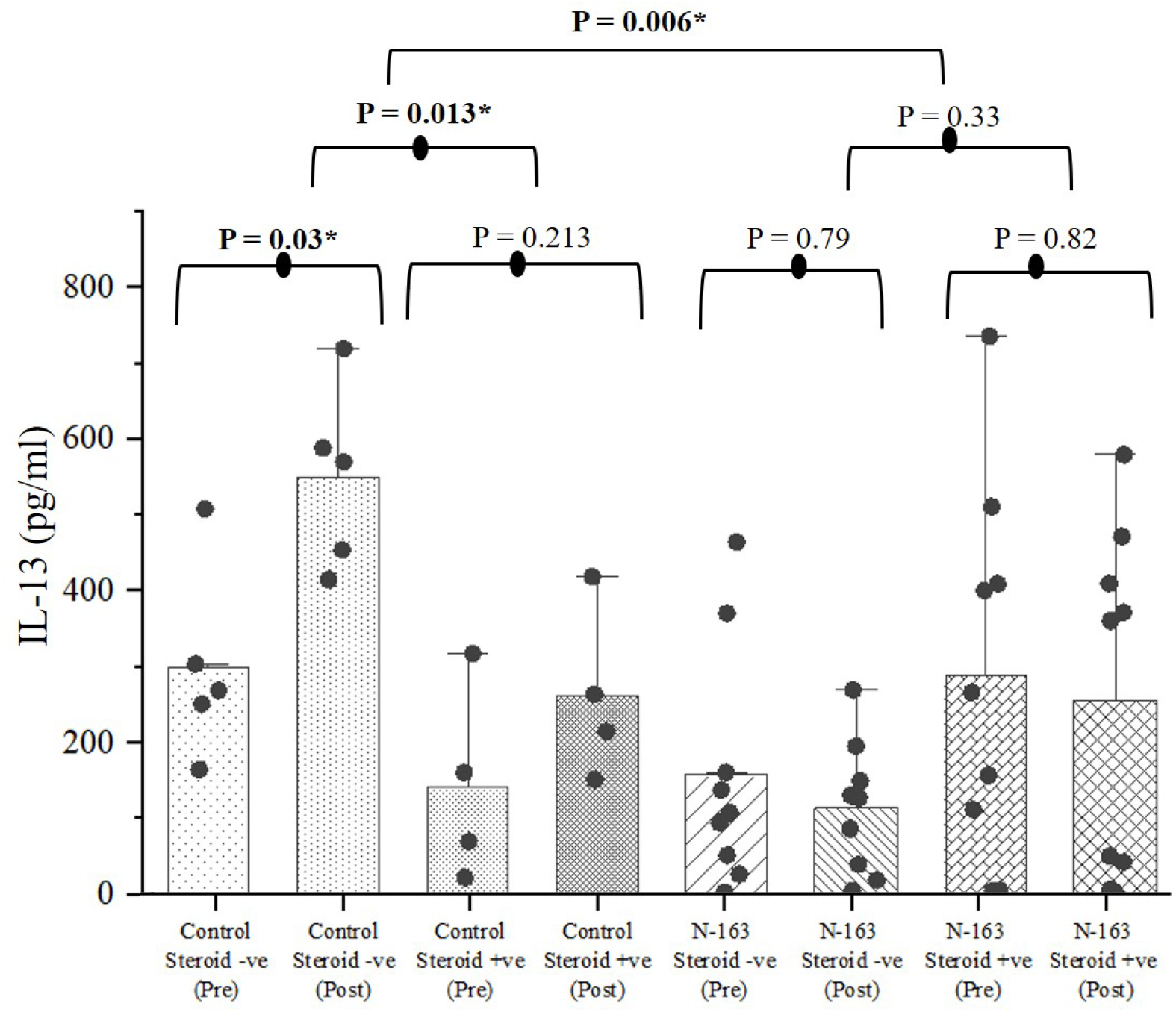
Levels of IL-13 levels showed statistically significant increase in control groups and decrease in treatment groups (*p-value significance < 0.05)

TGF-β levels showed a significant decrease in the N-163 Steroid -ve group, from a baseline value of 3302 ± 1895 pg/ml to 1325.66 ± 517 pg/ml post intervention, which was significantly lower than all the other groups (p-value = 0.02) (Figure 4, 6).

**Figure 4:**
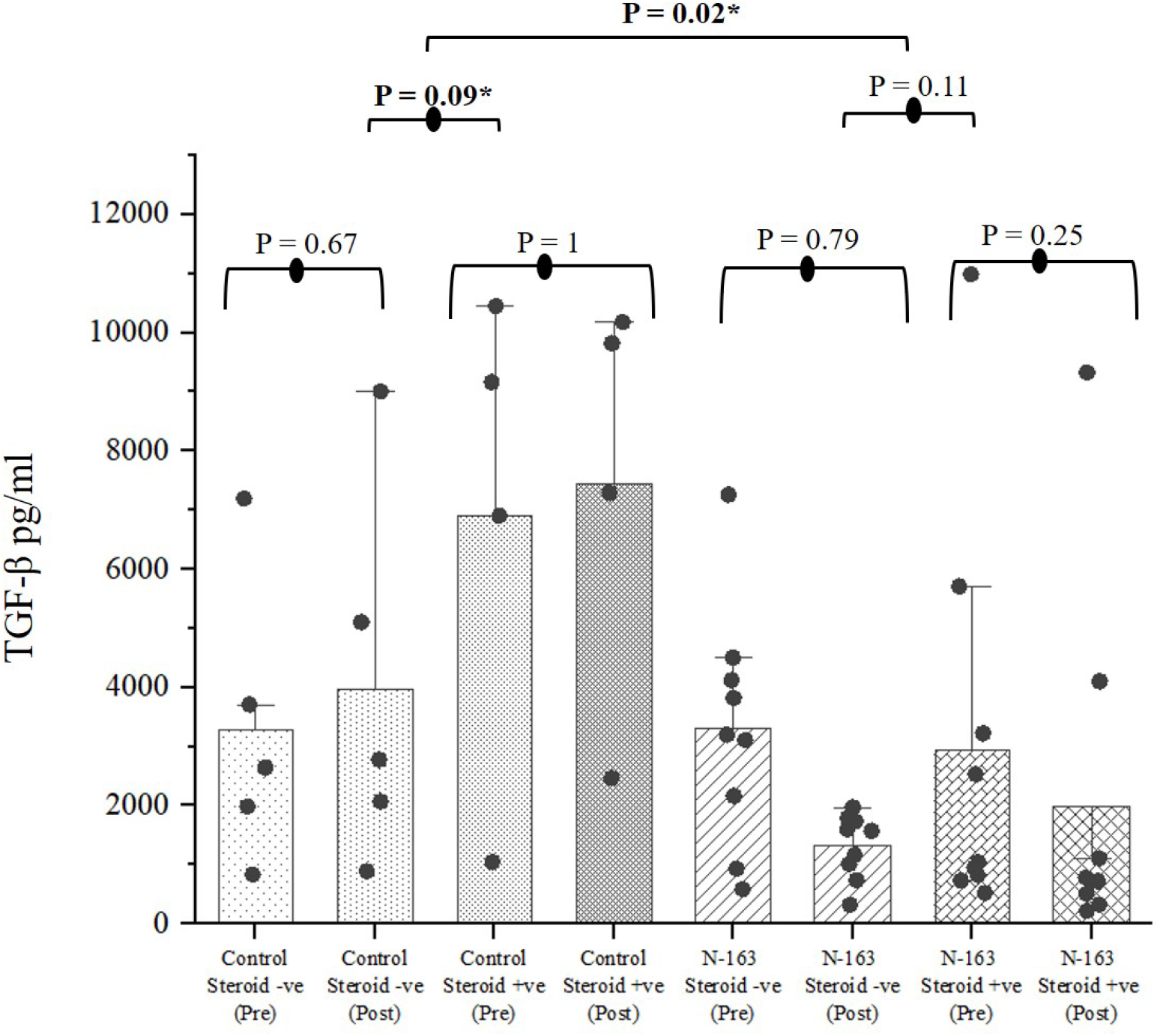
Levels of TGF-β showed significant decrease in the N-163 Steroid -ve group compared to other groups (*p-value significance < 0.05)

Dystrophin levels showed a significant increase in the N-163 Steroid -ve group, from a baseline value of 3.01 ± 1.58 ng/ml to 4.01 ± 1.44ng/ml post intervention, and the N-163 Steroid +ve group went from a baseline value of 3.15 ± 2.43 ng/ml to 3.78 ± 2.17 ng/ml post intervention, which was significantly higher than the control groups (p-value = 0.0009) (Figure 5, 6). The N-163 Steroid -ve group showed higher dystrophin expression than the N-163 Steroid +ve group with the difference being significant (p-value = 0.01). The percentage increase in dystrophin levels in the treatment group was up to 32.8%.

**Figure 5:**
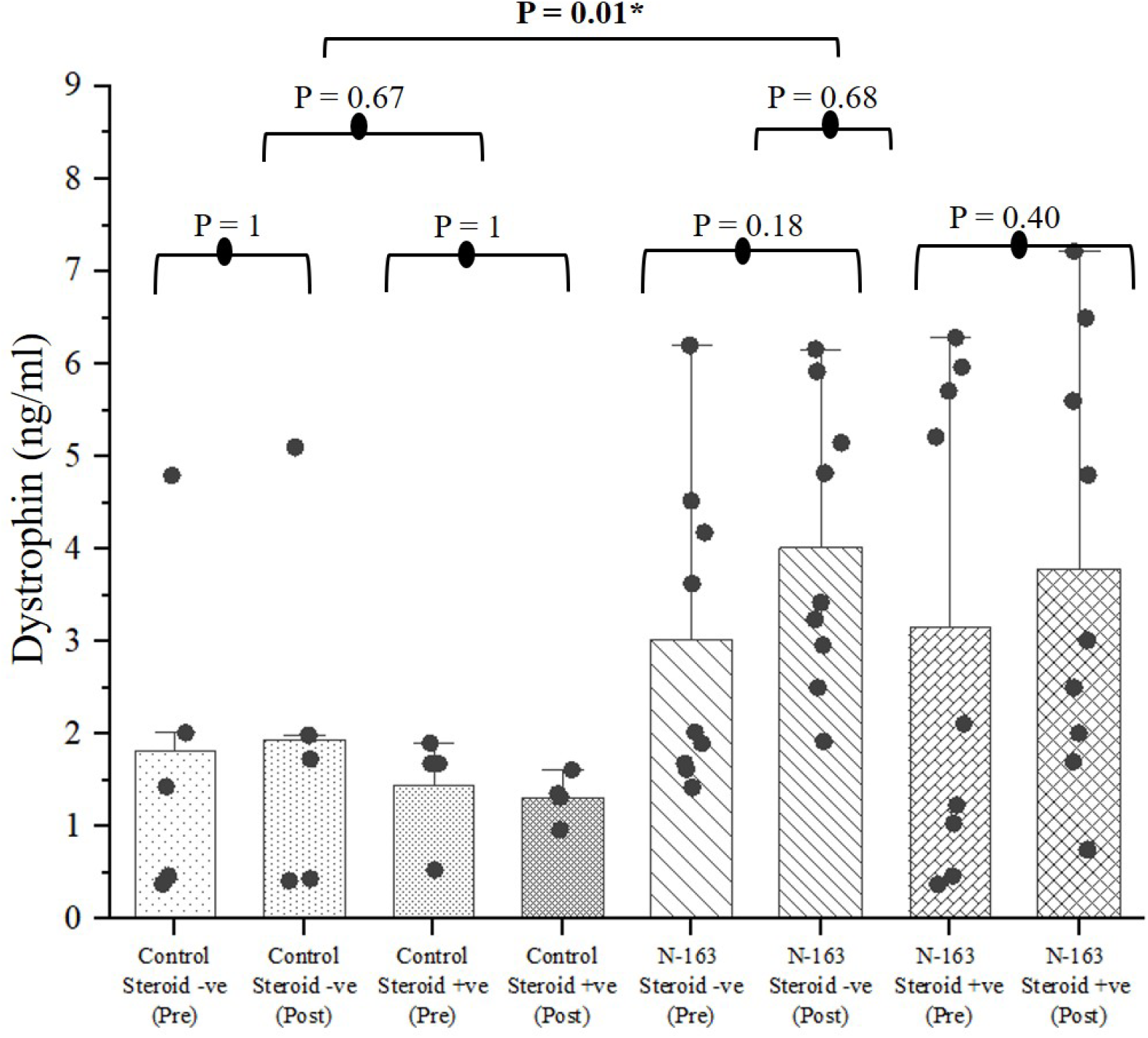
Levels of dystrophin showed significant increase in the N-163 Steroid -ve group compared to other groups (*p-value significance < 0.05)

Haptoglobin did not show much difference pre or post intervention in the treatment groups, but it was marginally increased in the control group (Figure 7). CK increased in the treatment groups (Figure 7). Urine myoglobin increased in the N-163 Steroid +ve group but decreased in all the other groups (Figure 7).

**Figure 6:**
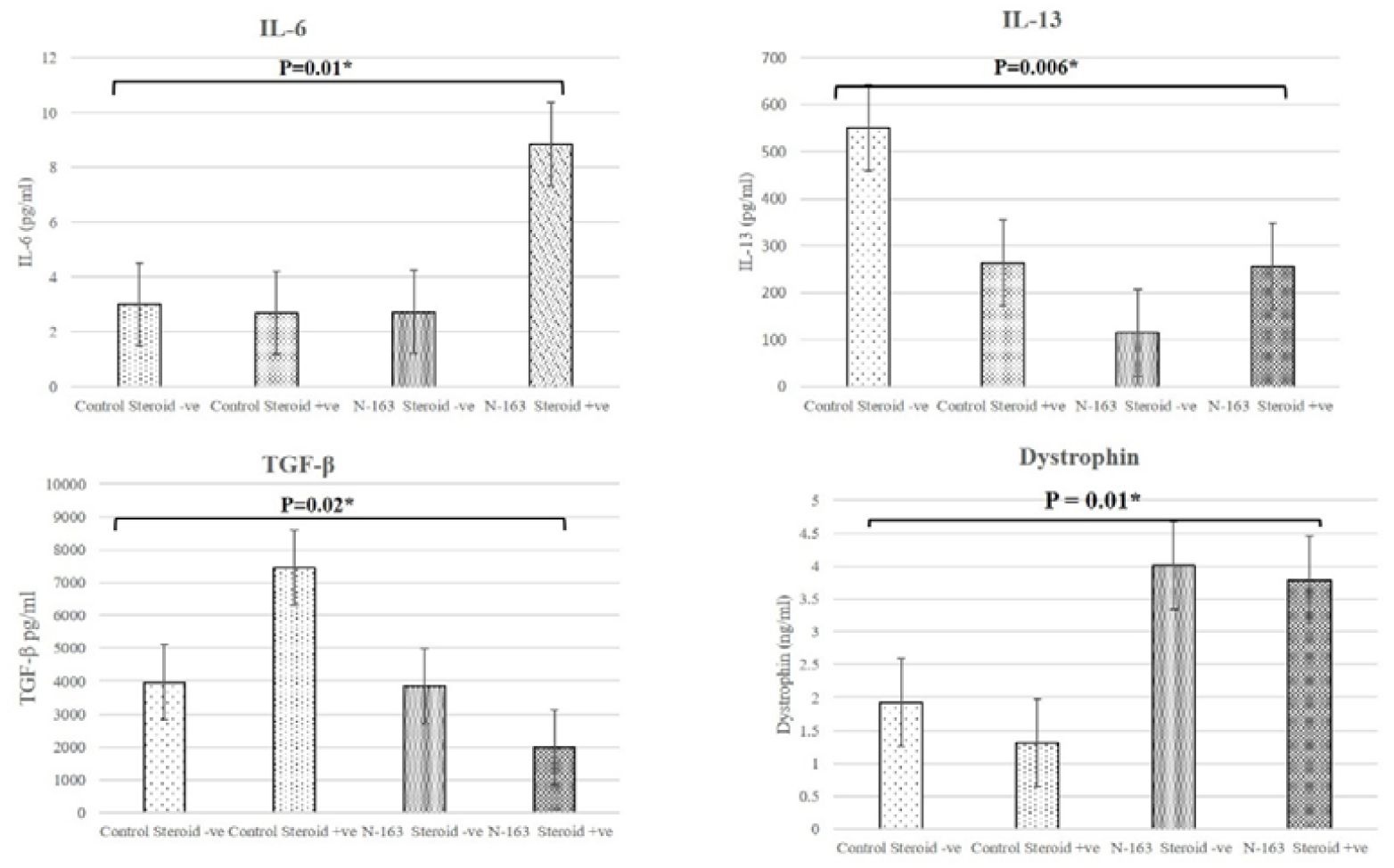
Comprehensive figure of IL-6, IL-13, TGF-β and Dystrophin showing comparison of values between control and treatment group (N-163), pre- and post-intervention (*p-value significance < 0.05)

**Figure 7:**
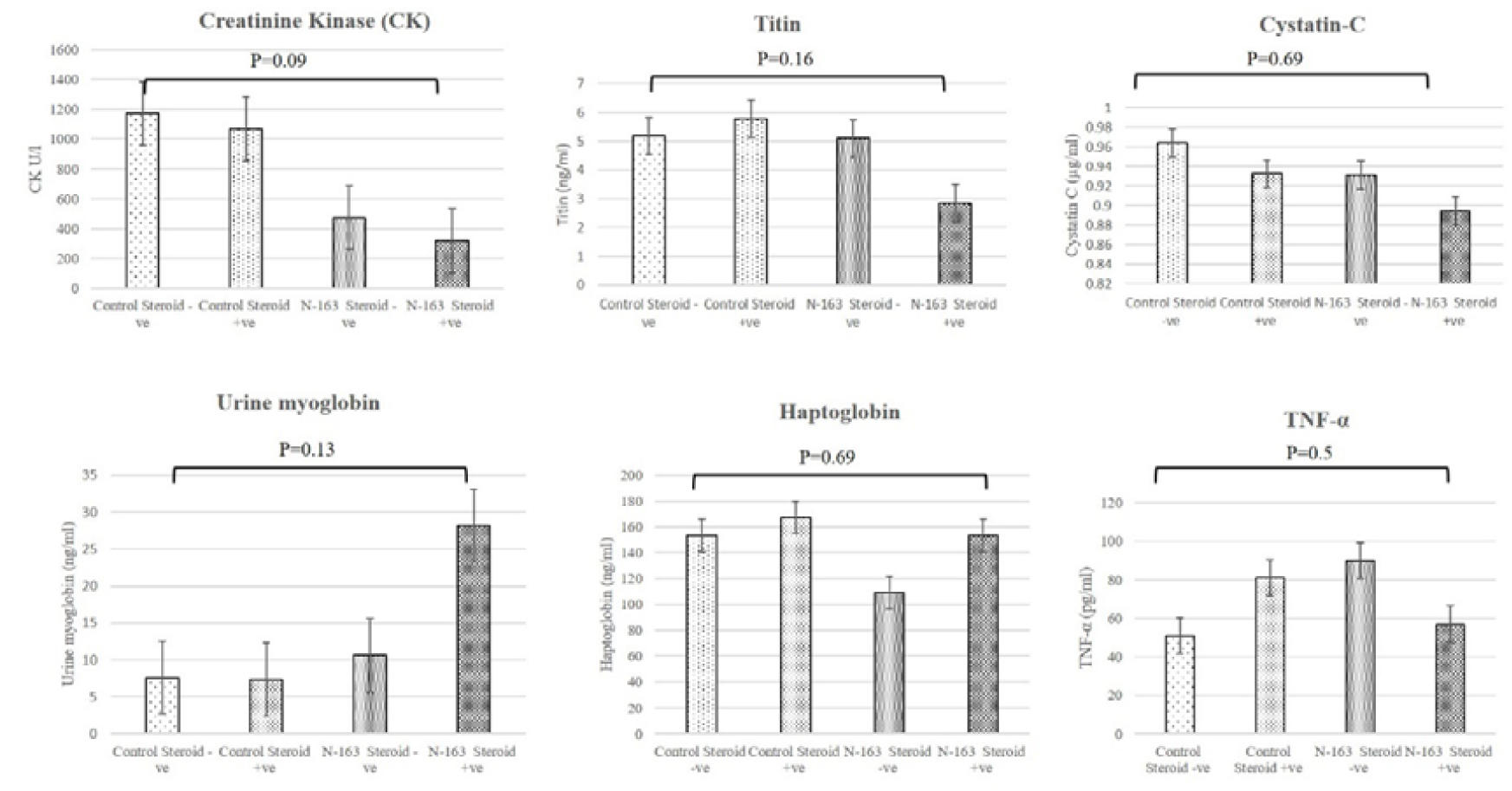
Levels of Creatinine kinase (CK), Titin, Cystatic-C, Urine myoglobin, Haptoglobin and TNF-α between control and treatment group (N-163), pre- and post-intervention (*p-value significance < 0.05)

Titin and cystatin C decreased in the N-163 Steroid +ve group and the control Steroid +ve group, but the difference was not significant. TNF-α decreased in all the groups except the N-163 Steroid -ve group (Figure 7).

The 6MWT and NSAA did not show any significant differences between the groups (Figure 8). The MRC grading showed slight improvement in 12 out of 18 patients (67%) in the treatment group and only four out of nine (44%) subjects in the control group but it was not statistically significant (p-value = 0.02) (Table 3, Figure 8).

**Table 3:** Medical research council (MRC) grading of muscle power in the study groups

| Subject | Ambulatory/Non-ambulatory | Baseline | Post-intervention | Progression |
| --- | --- | --- | --- | --- |
| **Control group** |  |  |  |  |
| 1 | Ambulatory | 151 | 165 | Improved |
| 2 | Ambulatory | 133 | 162 | Improved |
| 3 | Non-ambulatory | 70 | 80 | Improved |
| 4 | Non-ambulatory | 164 | 164 | No change |
| 5 | Non-ambulatory | 109 | 106 | Worsened |
| 6 | Non-ambulatory | 102 | 105 | Improved |
| 7 | Non-ambulatory | 117 | 115 | Worsened |
| 8 | Non-ambulatory | 110 | 109 | No change |
| 9 | Non-ambulatory | 105 | 102 | Worsened |
| **Treatment Group** |  |  |  |  |
| 1 | Ambulatory | 134 | 133 | Worsened |
| 2 | Ambulatory | 146 | 163 | Improved |
| 3 | Ambulatory | 119 | 131 | Improved |
| 4 | Ambulatory | 127 | 136 | Improved |
| 5 | Ambulatory | 154 | 160 | Improved |
| 6 | Ambulatory | 158 | 168 | Improved |
| 7 | Non-ambulatory | 102 | 119 | Improved |
| 8 | Non-ambulatory | 116 | 117 | Improved |
| 9 | Non-ambulatory | 127 | 127 | No change |
| 10 | Non-ambulatory | 120 | 131 | Improved |
| 11 | Non-ambulatory | 96 | 107 | Improved |
| 12 | Non-ambulatory | 111 | 122 | Improved |
| 13 | Non-ambulatory | 117 | 113 | Worsened |
| 14 | Non-ambulatory | 108 | 108 | No change |
| 15 | Non-ambulatory | 103 | 107 | Improved |
| 16 | Non-ambulatory | 145 | 148 | Improved |
| 17 | Non-ambulatory | 93 | 119 | Improved |
| 18 | Non-ambulatory | Test couldn't be performed |  |  |

**Figure 8:**
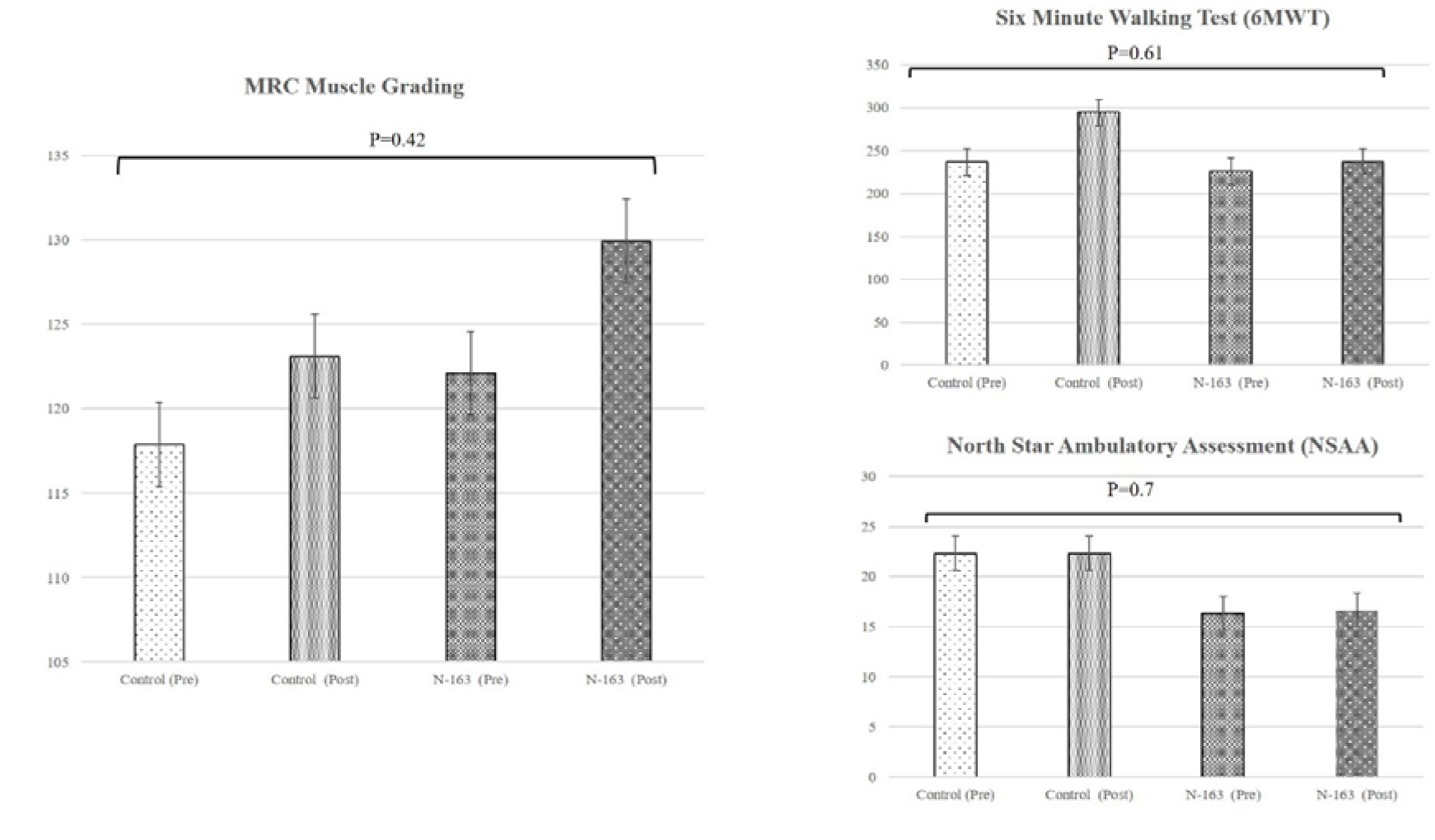
MRC muscle grading, 6MWT and NSAA results between control and treatment group (N-163), pre- and post-intervention (*p-value significance < 0.05)

## Discussion

Current interventions for DMD, such as corticosteroids and rehabilitative care, help to prolong survival up to the third or the fourth decade of life. Corticosteroids remain the mainstream supportive approach to slow inflammation and the associated decline in muscle strength and function [4]. However, steroids have their own adverse effects, and their prescription is based on risk versus benefit to that specific patient and tolerance to the medication. Exon-skipping gene therapy and cell-based strategies to replace the mutant DMD gene are in development, but the desired outcome has not yet been achieved. In the meantime, nutraceuticals can be considered potential strategies for immune modulation and alleviating inflammation, as they are safer with lesser adverse effects [4]. Improvement of the locomotor performances and mitochondrial respiration by 1,3-1,6 beta-glucans in zebra fish model of muscular dystrophy [6] has already been reported.

In the current study, we focussed on a 1-3,1-6 beta glucan from the N-163 strain of the black yeast A. pullulans that has been reported to mitigate inflammation, evidenced by a decrease in anti-inflammatory markers and production of beneficial immuno-modulation [6-8]. The safety profile of N-163 beta glucan has been confirmed by the results.

### Anti-inflammatory and anti-fibrotic outcomes

Circulating IL-6 is chronically elevated in individuals with DMD [13], which has been reported to contribute to DMD-associated cognitive dysfunction. IL-6 blockades have been advocated as a therapeutic approach for DMD [14]. In the present study, IL-6 showed highest decrease in the N-163 Steroid -ve group (Figure 2). While IL-6 is an acute inflammatory biomarker [14], IL-13 is a pro-fibrotic biomarker [15] and was significantly decreased (Figure 3). Together with the TGF-β pathway, it is a major proinflammatory and pro-fibrotic cytokine responsible for the chronic inflammatory response leading to replacement of the muscle by scar tissue or fibrosis, resulting in muscle weakness and loss of muscle function [16]. IL-6 values matched that of healthy controls and DMD patients who were treated with corticosteroids [17]. TGF-β levels also showed a significant decrease in the N-163 Steroid -ve group (Figure 4). Dystrophin restoration of 20% expression [18,19] is considered the point of efficacy for a DMD therapy [20] and was found to increase by 32.8% in both the treatment groups (Figure 5) of the present study from baseline. We used serum dystrophin evaluation as recent evidence points to the fact that the lack of dystrophin in the smooth muscle of blood vessels is the major contributing factor to skeletal muscle pathogenesis in DMD and not the dystrophin deficiency in skeletal or cardiac muscle [21]. Dystrophin is expressed in the tunica media of blood vessels, especially in the muscular arteries. It has also been reported that the blood vessel-associated dystrophin is encoded by another gene than the skeletal muscle dystrophin gene [22]. In a mdx mice model study, it was shown that dystrophin which was absent in the blood vessels could be restored by induced genetic expression of full-length human dystrophin (h-dys) and this expression was able to restore the NO-dependent attenuation of α-adrenergic vasoconstriction in the exercising muscles, preventing further exacerbation of muscle fibre necrosis [23]. In the present study, an increase in the plasma levels of dystrophin, which must have been derived from vascular smooth muscle is evidence for the efficacy of beta glucans as a potential adjunct to restore dystrophin in DMD.

### Other biochemical markers of relevance

While haptoglobin and urine myoglobin did not show significant differences, the increase in urine myoglobin in the N-163 Steroid +ve group deserves more analysis concerning the underlying mechanism. Greater activity among steroid-treated individuals may place their dystrophin-deficient muscles under greater mechanical stress, predisposing them to further muscle fibre damage and consequent myoglobinuria [24]. While titin and cystatin C decreased in the N-163 steroid +ve group and in the control Steroid +ve group, there was an increase in CK, which is paradoxical, as reports suggest that titin concentration correlates significantly with serum CK concentration [25].

### Muscle strength evaluation

There were three evaluations to assess muscle strength and tone, done in a blinded manner by the same physiotherapist at baseline and post intervention. Though the 6MWT and NSAA did not show any significant differences between the groups, MRC grading showed improvement of muscle strength in 67% of the subjects in the treatment group compared to 44% subjects in the control group, which is significant. The limitation of this being a 45-day study is relevant to the muscle-strength and functional evaluations, mandating the need for a longer study and follow-up duration. However, though small, the improvement in MRC grading at 45 days could be again attributed to the immune modulation effects of this disease-modifying supplement. The study shows proof of concept that DMD could be tackled by the N-163 beta glucan from three aspects: decrease in inflammation shown by decreased IL-6 and TNF-α, decrease in fibrosis evident by decreased TGF-β and IL-13 and, more importantly, restoration of dystrophin evident from a 32.8% increase in dystrophin levels. These effects hold regardless of the use or non-use of steroids, which is important, as this safety-proven food supplement can help DMD patients regardless of steroid status.

Chronic inflammation being common to pathogenesis of all muscular dystrophies, immunomodulatory treatment may benefit patients with diverse types of muscular dystrophy [26]. Further, modulating the inflammatory response and inducing immunological tolerance to de novo dystrophin expression is critical to the success of dystrophin-replacement therapies [27]. The need to evaluate the muscles involved in respiratory function and myocardium should be mentioned here, as they are the cause of mortality in most of the DMD patients [1]. Though other dystrophies, such as limb girdle muscular dystrophy (LGMD), do not involve respiratory or cardiac muscles, inflammatory overactivity is the common pathophysiology among types of muscular dystrophy [26]. Once proven efficacious for DMD, extending the beneficial application of the N-163 beta glucans to other dystrophies such as LGMD can be considered.

DMD is a rare genetic disease with a maximum life expectancy of up to fourth decade, with the majority of victims dying in their late twenties to thirties. The average lifespan at birth, which was 20+ years for those born in or before 1970, has gradually increased by 10∼15 years for those born and diagnosed with DMD in the 1980s and 1990s. This increase is attributed to better or early ventilatory assistance, steroid usage and cardiac care [28,29], which are only supportive interventions. With the gene therapies approved recently, there is a hope of additional progress and increase in lifespan [30]. Though these gene therapies (such as exon skipping) address the root cause by splicing out selected exons from the pre-mRNA at or next to the mutation site, generating a translatable transcript from the mutant dystrophin gene leading to dystrophin expression [30, 31], they are still marred by challenges such as delivery of gene-editing components throughout the musculature and mitigation of possible immune responses [32]. The current need, therefore, is to modulate the immune system and control the inflammation and ensuing fibrosis to delay the progression of the disease. The earlier usage of steroids in a regular manner was later changed to intermittent usage [33] with regimens varying between institutes; now, newer steroids with lesser adverse effects are in various stages of progress towards clinical applications [34]. In this background, the safety of this N-163-produced beta glucan food supplement without adverse reactions is to be considered an indispensable value addition. Targeting the inflammation component (the criteria for selecting this supplement for this study) having yielded beneficial outcomes, additional studies on this characteristic could be of value to possibly extending their application for other neuroinflammatory diseases, such as multiple sclerosis.

The limitations of the study include uneven distribution of subjects, broad age range (5-19 years) and short follow-up (only 45 days); improvements in muscle function over the course of the study showed variability that may have been due to the level of sensitivity to change of functional assessments during the disease progression in the age group. Among the 27 subjects, two-thirds were ambulatory and the remaining non-ambulatory; the evaluation criteria differences must be kept in mind, which may show equivalent quantification among all DMD patients at different stages of disease severity when non-invasive myograms to measure the individual muscles accurately could be undertaken. Further, the steroid dose was not uniform and fell into a broad range (6-24 mg). Also, the consumption of steroids vs those who did not consume them or those who had stopped steroids after an initial duration of consumption, as well as regimen variation, are to be considered while interpreting the outcomes. All these aspects mandate the need for larger randomized clinical trials of longer duration to validate this supplement as a treatment.

## Conclusion

N-163 beta glucan with and without steroids helped decrease IL-6, TGF-β and IL-13 and increase dystrophin levels along with improvement of muscle strength in subjects with DMD in this clinical study. Thus, N-163 beta glucan is a safe and effective potential therapeutic adjunct for patients with DMD. While the benefits documented may help slow the rate of progression of this devastating disease, confirmation by longer and larger studies will help establish this agent for routine clinical application as a disease-modifying agent with the potential to help prolong the lifespan of DMD patients. After such validation, extending its application to other dystrophies such as LGMD could be considered, and further in-depth research in neuroinflammatory diseases are likely to shed light on the mechanism of action, leading to additional beneficial applications.

## Data Availability

All data produced in the present work are contained in the manuscript

## Declarations

### Ethics approval and consent to participate

The study was registered in Clinical trials registry of India, CTRI/2021/05/033346. Registered on 5th May, 2021. The study was approved by the Institutional Ethics Committee (IEC) of Saravana Multispeciality Hospital, India on 12th April, 2021.

### Consent for publication

Not applicable

### Availability of data and material

All data generated or analysed during this study are included in the article itself.

### Authors’ contributions

KR, NI and SA. contributed to conception and design of the study. KR, RS and NI helped in data collection and analysis. SA and SP drafted the manuscript. VD, SS, SUP, SSU and MI performed critical revision of the manuscript. All authors read and approved the final manuscript.

## Acknowledgements

The authors would like to dedicate this paper to the memory of Mr. Takashi Onaka, who passed away on the 1st of June, 2022 at the age of 90 years, who played an instrumental role in successfully culturing an industrial scale up of AFO-202 and N-163 strains of Aureobasidium pullulans after their isolation by Prof. Noboru Fujii, producing the novel beta glucan described in this study.

They thank

1. The Government of Japan and the Prefectural Government of Yamanashi for a special loan and M/s Yamanashi Chuo Bank for processing the transactions.
2. Ms. Sunitha, Mr. Vincent, Mr. Shivakumar (Physiotherapist), Dr. Madhankumar and the staff of Kenmax & Sarvee Integra, for their assistance during the clinical study and data collection of the manuscript.
3. Fr. Francis Xavier, Fr. Vargheesh Antony, Fr. Marianathan and Mr Jeyachandran of JAICARE for their support during the clinical study.
4. Mr. Mitsuru Nagataki (Sophy Inc, Kochi, Japan), for necessary technical clarifications.
5. Mr. Yoshio Morozumi and Ms. Yoshiko Amikura of GN Corporation, Japan for their liaison assistance with the conduct of the study.
6. Loyola-ICAM College of Engineering and Technology (LICET) for their support to our research work.

## Abbreviations

DMD: Duchenne muscular dystrophy
CK: Creatinine kinase
TNF-α: tumour necrosis factor – Alpha
MRC: Medical research council
NLR: Neutrophil-to-lymphocyte ratio
LCR: Lymphocyte-to-CRP ratio
LeCR: Leukocyte-to-CRP ratio
NASH: Non-alcoholic steatohepatitis
6MWT: Six-minute walk test
NSAA: North Star Ambulatory Assessment
LGMD: Limb girdle muscular dystrophy

